# Geographic access to COVID-19 healthcare in Brazil using a balanced float catchment area approach

**DOI:** 10.1101/2020.07.17.20156505

**Authors:** Rafael H. M. Pereira, Carlos Kauê Vieira Braga, Luciana Mendes Servo, Bernardo Serra, Pedro Amaral, Nelson Gouveia, Antonio Paez

## Abstract

The rapid spread of the new coronavirus across the world has raised concerns about the responsiveness of cities and healthcare systems during pandemics. Recent studies try to model how the number of COVID-19 infections will likely grow and impact the demand for hospitalization services at national and regional levels. However, less attention has been paid to the geographic access to COVID-19 healthcare services and to the response capacity of hospitals at the local level, particularly in urban areas in the Global South. This paper shows how transport accessibility analysis can provide actionable information to help improve healthcare coverage and responsiveness. It analyzes accessibility to COVID-19 healthcare at high spatial resolution in the 20 largest cities of Brazil. Using network-distance metrics, we estimate the vulnerable population living in areas with poor access to healthcare facilities that could either screen or hospitalize COVID-19 patients. We then use a new balanced floating catchment area (BFCA) indicator to estimate spatial, income and racial inequalities in access to hospitals with intensive care unit (ICU) beds and mechanical ventilators while taking into account congestion effects. Based on this analysis, we identify substantial social and spatial inequalities in access to health services during the pandemic. The availability of ICU equipment varies considerably between cities and it is substantially lower among black and poor communities. The study maps territorial inequalities in healthcare access and reflects on different policy lessons that can be learned for other countries based on the Brazilian case.

## 1. Introduction

The global outbreak of the new coronavirus (SARS-CoV-2) has raised serious concerns about the responsiveness of healthcare systems and particularly about how vulnerable population groups might be affected (Lancet, 2020; WHO, 2020). A rapidly growing body of research has emerged to model how the number of COVID-19 infections will likely grow and impact the demand for hospitalization services globally (Petropoulos & Makridakis, 2020; Walter et al, 2020) and at the national level (Arenas et al., 2020; Moghadas et al., 2020; Paez, 2020; Paez et al., n.d.; Wu, Leung, & Leung, 2020). However, less attention has been paid to the geographic access to COVID-19 healthcare services and to the response capacity of hospitals at the local level in urban areas, despite the potential relationships between accessibility to healthcare resources and mortality (Ji, Ma, Peppelenbosch, & Pan, 2020). Early work by Ji et al. (2020) and Rader et al. (2020), for example, considered resources at the provincial level in China and at the county level in the USA, but we are not aware of studies that investigate the issue of resource allocation at higher spatial resolutions, particularly in the context of Latin America, where the epicenter of the pandemic shifted in June, 2020.

The goal of this study is to present estimates of geographic accessibility to COVID-19 healthcare at high spatial resolution in the 20 largest cities of Brazil. Healthcare services in Brazil are known to be unevenly distributed across the country and also within cities (Amaral et al., 2017). In this context, it is crucial to map where vulnerable social groups confront poor accessibility to health services. Similarly, it becomes paramount to identify which healthcare facilities are likely to face surges in demand due to the need to hospitalize severely ill patients. In this paper we combine traditional and novel accessibility metrics to address these questions. Using network-distance metrics, we first estimate the number of vulnerable people living in areas with poor access to inpatient or outpatient facilities able to provide care for patients with suspected or confirmed cases of COVID-19. Next, we use a new balanced floating catchment area method (BFCA) proposed by (Paez, Higgins, & Vivona, 2019) to analyze levels of access to hospitals that could treat patients with severe symptoms of COVID-19, taking into account healthcare system capacity and competition effects for ICU beds with mechanical ventilators.

The results of this research are useful to identify, in the 20 cities examined, a population of approximately 1.6 million people who live more than 5 km away from a healthcare facility equipped to treat severe cases of COVID-19. Furthermore, although overall the average number of intensive care units (ICU)beds with ventilators is 1.06 per 10 thousand inhabitants, there are large variations in this level of service, both between and within cities. In particular, we find that the accessibility to ICU resources is substantially lower in black and poor communities. This creates a worrying scenario given the strong potential for propagation of COVID-19 combined with poor health outcomes. The study maps territorial inequalities in healthcare access and reflects on different policies that could be adopted to address them.

The remainder of this paper is as follows. The next section provides relevant background information regarding the evolution of the COVID-19 pandemic in Brazil and accessibility analysis. This is followed by a discussion of the data and methods used in this research. Then, the results of the empirical analysis are presented and discussed. And finally, we offer some concluding remarks, including policy implications and directions for future research.

## 2. Background

### 2.1 COVID-19 in Brazil

The first confirmed case of COVID-19 in Latin America was in late February 2020, in Brazil. The affected person was a man from São Paulo who had travelled that month to Italy. This case was typical of the beginning of the epidemic in Brazil, with other early cases of the disease imported via international flights coming mostly from Italy and the United States (Candido et al., 2020). At that early stage, the then-Minister of Health noted that it would remain to be seen how the virus behaved in a tropical country in the middle of summer. While there is evidence that incidence of the disease is lower at higher temperatures (Paez et al., 2020), it is now clear that the disease can be devastating during summer too: as early of July, less than five months after the first confirmed case of the COVID-19 in Brazil, over 1.8 million cases have been confirmed, and over 70 thousand deaths have been attributed to the disease, 45% of which concentrated in the 20 largest cities of the country. By early June, Brazil had become, after the United States, the country with the highest number of cases of COVID-19 in the world.

The earliest cases of COVID-19 in Brazil were concentrated among middle and upper-class people (Souza et al., 2020). In Brazil’s mixed health care system, these segments of the population typically can afford to pay for healthcare or use health services intermediated by private health insurance. This is not the case for lower-income groups, who are largely dependent on the public health system in Brazil, and among whom community transmission rapidly increased the number of infections. This development is particularly worrisome as low-income groups in the country also typically live in less developed urban areas with poor transport services and poor access to health, education, and employment opportunities (Pereira, Braga, Serra, & Nadalin, 2019). Previous research, in fact, has identified important spatial gaps in accessibility to emergency services in Brazil (Rocha et al., 2017). To further complicate matters, other research has linked poor accessibility to higher pneumonia mortality (Zaman et al., 2014).

Given the rapid growth of COVID-19 in Brazil, it is important to map the potential stress on the country’s healthcare system. Preliminary reports already show an unusual increase in the numbers of admissions to hospitals due to suspected COVID-19 infections (InfoGripe, 2020). These studies raise serious concerns about the overload the pandemic can generate to the public Unified Health System (Sistema Único de Saúde - SUS), which already started showing signs of collapse similar to those observed in Italy and Spain (Grasselli, Pesenti, & Cecconi, 2020; Legido-Quigley et al., 2020). The work of Noronha et al. (2020), for example, presented a simulation of nationwide spread of the new coronavirus and analyzed the supply and demand for ICU beds with ventilators, broken down by health regions. They found that even in an optimistic scenario, of an infection level of 0.1% in the first month, roughly half of the nation’s health regions would face a grave deficit of ICU beds to meet demand for admission of patients with severe cases of COVID-19. Similar regional modeling studies conducted by Coelho et al (2020) and Castro et al (2020) suggest the pressure on the health system is more likely to reach critical levels in large urban centers, where the number of confirmed cases is higher.

In this context, there is still a lack of studies that look at the COVID-19 healthcare provision at the city level, particularly at what actionable insights can be drawn from analysis of vulnerable groups and their access to health services in high spatial resolution. The looming crisis faced by the health system due to the COVID-19 requires many emergency actions. For this purpose, it is essential for healthcare planners to have a diagnosis of the areas of cities with less access to health services and equipment, and to identify the hospitals that might suffer overloaded demand for admissions.

### 2.2 Healthcare accessibility

One of the most commonly used indicators to measure geographic access to healthcare is the shortest distance/travel time to the closest facility (Geurs & van Wee, 2004; Neutens, 2015). This indicator is widely used in part because it is relatively simple to calculate and straightforward to interpret, and thus easily communicated to policy makers. A well-known limitation of this indicator, however, is that it overlooks competition effects since it does not account for potential population demand nor for the levels of service supply.

Another popular approach to measure access to healthcare is the family of Floating Catchment Area (FCA) methods (Matthews et al., 2019). A key advantage of this family of indicators is that it accounts for capacity restrictions, local competition effects as well as cross-border healthcare-seeking behavior (Neutens, 2015). The common rationale underlying FCA methods is to calculate accessibility levels in sequential steps. This first step is to calculate the provider-to-population ratio (PPR) of each health facility as a ratio between its service supply (e.g. number of ICU beds) and its potential service demand given by the population that falls within some catchment area. The second step is to calculate accessibility levels of each population center by aggregating the PPR of every healthcare provider that is accessible from each population center.

The first indicator of this sort is the two-step floating catchment area (2SFCA), proposed in the early 2000s (Luo & Wang, 2003; Radke & Mu, 2000). Since then, multiple authors have proposed incremental improvements to the basic model in order to incorporate more sophisticated impedance functions (Dai, 2010; Luo & Qi, 2009), to consider suboptimal configurations of health systems (Delamater, 2013) and to account for spatially adaptive floating catchments (Matthews et al., 2019; Matthew R. McGrail & Humphreys, 2009) and trip-chaining behavior (Fransen, Neutens, De Maeyer, & Deruyter, 2015).

A fundamental limitation of FCA methods is that they overestimate both service demand and supply, which can generate misleading accessibility estimates (Delamater, 2013; Paez et al., 2019; Wan, Zou, & Sternberg, 2012). Demand inflation occurs when populations that fall within the overlap of catchment areas are counted more than once as potential demand for multiple facilities. Supply inflation, on the other hand, happens when the level of service of a healthcare unit is simultaneously allocated to multiple population centers (Paez et al., 2019). Until recently, two approaches had been proposed to address this issue. Wan et al. (2012) introduced the Three-Step Float Catchment Area (3SFCA), which deflates demand by introducing an initial step that splits the potential demand of a population center over multiple health facilities proportional to transport costs/distances. Meanwhile, Delamater (2013) proposed the Modified Two-Step Floating Catchment Area (M2SFCA), which deflates the supply side by increasing the friction of distance in a way that allocates levels of service more locally. Nonetheless, both methods only partially fix the inflation problem, as they compound the effects of impedance functions to address either demand inflation (3SFCA) or supply inflation (M2SFCA).

To overcome this limitation, Paez et al. (2019) recently introduced a new indicator to the FCA family, which we term here as the Balanced Float Catchment Area (BFCA). The new BFCA uses a standardized impedance matrix to generate proportional allocation of demand and level of service, fixing both demand and supply inflation issues in FCA calculations. The result is a more intuitive measure of accessibility that 1) accounts for competition effects, 2) provides a local version of the provider-to-population ratio (PPR) that is interpreted similarly to a regional PPR; and 3) preserves system-wide population and level of service, overcoming inflation issues. Estimating accessibility during a pandemic is an idoneous application of this method because congestion over the short term is one of the fundamental issues to address. Therefore the analysis must account accurately for competition for scarce resources, if policy interventions are to have any hope of addressing shortfalls effectively.

## 4. Data and Methods

### 4.1 Data

Accessibility to health facilities is estimated using data generated by the Access to Opportunities Project (Pereira et al, 2019)^1^. The method combines data from national household surveys, administrative records of the federal and municipal governments, along with satellite images and collaborative mapping data to estimate accessibility at high spatial resolution on a hexagonal grid with size of 357 meters (short diagonal), approximately the size of a typical city block.

Original sociodemographic data comes from the 2010 population census conducted by the Brazilian Institute of Geography and Statistics (IBGE). These data are aggregated in an hexagonal grid using dasymetric interpolation in two steps, as follows. Data on population count, income, race and age distribution are gathered at the census tract level. Population counts are then updated based on municipal-level demographic projections for 2020, published by Freire et al. (2020). The total projected population of each city in 2020 was distributed across census tracts assuming that the relative distribution of the population by district and age cohort of each sector remained constant between 2010 and 2020. We then use dasymetric interpolation to pass information from census tracts to a finer regular grid of 200 meters with population count data considering aerial intersection and population sizes. Finally, these data are reaggregated from the regular grid to the hexagonal grid.

Data on healthcare facilities associated with the SUS, as registered in the National Registry of Health Facilities (CNES), at the end of 2019 were geocoded and made publicly available by Pereira et al. (2019). For this paper, these data were complemented with updated information from the CNES for February 2020 about the number of adult ICU beds and ventilators in each healthcare facility. We also included geolocated data on 30 field hospitals and reactivated hospitals in 15 cities up to April 2, 2020. The functioning of these hospitals has eased demand on other hospitals and expanded the capillarity of the health system, by adding a total of 868 beds in ICUs, ITCs and semi-intensive or semi-critical care units. Finally, we used collaborative mapping data from OpenStreetMap and data on terrain from satellite images (JAXA, 2011). These data were processed with OpenTripPlanner, an open routing algorithm for multimodal transport networks, to generate the door-to-door travel times between all the hexagon pairs in each city.

### 4.2 Methods

The analysis presented in the paper is divided in two parts. In the first part, we estimate for Brazil’s 20 largest cities the number and living places of vulnerable populations who: (a) cannot access within 30 minutes on foot an establishment associated with the SUS that can perform triage and refer patients suspected of COVID-19 infection for hospitalization; and (b) live further than 5 Km from a hospital with capacity to admit patients suffering from SARS with adult ICU beds and ventilators. Vulnerable population groups were defined as people above 50 years old with lower income (in the bottom half of the income distribution). This criteria was chosen because the resulting group includes people who: 1) are at greater vulnerability to COVID-19 infection due to their age; 2) are more dependent on the public health system; and 3) tend to face greater difficulties of urban mobility and access to health services^2^.

All time and distance thresholds were chosen as a first exploratory analysis following the official recommendation from the Health Ministry and local officials (state and municipal), who recommend that people with suspected COVID-19 infection stay at home if the symptoms are mild or go to the nearest health unit for an initial interview (anamnesis) and notification of the surveillance team (Brasil, 2020b). Patients with severe symptoms should be referred for admission to hospitals in general wards or ICUs (ibid.). In Brazil, the typical pattern is for people to use hospitals as entry points for daily health services and emergency care (ibid). Following clinical management and fast track service protocols (Brasil, 2020a, 2020b), this analysis included those primary health service facilities with capabilities to screen patients suspected of COVID-19, referer patients to specialized services, as well as facilities with more advanced service levels, such as emergency care centers, first aid posts and hospitals^3^.

Accessibility levels by public transportation were not considered because the use of collective transportation is not recommended for people with symptoms of COVID-19. Besides this, various cities saw a drop of over 70% in the supply of transit services as a response to isolation measures (NTU, 2020). This reduction makes public transport services less reliable and also more hazardous for contagion due to the large agglomeration of people at stops/stations and aboard vehicles.

In the second part of the analysis we look at access to health services while taking into account healthcare system capacity and competition effects. In this part we focus on access to those facilities that could provide hospitalization in ICUs to support patients with COVID-19. This analysis estimates provider-to-population ratios from both origin and destination perspectives. We calculated (1) for each hexagonal cell the number of accessible ICU beds with ventilators, and (2) for each hospital the ratio between the number of adult ICU beds with ventilators available and the population of the corresponding catchment area.

Here we use the balanced float catchment area (BFCA) to calculate component (1) and a partial version using the first two steps of BFCA to calculate component (2). There are generally two approaches to implement accessibility measures: positive and normative. The first one considers the willingness of people to travel whereas the latter captures a norm to be satisfied (Páez, Scott, & Morency, 2012). In practice, the difference between positive and normative accessibility is the definition of the impedance function. In this study we consider both approaches. In the first part of the empirical analysis we consider a normative implementation with a threshold of 30 minutes travel time based on research by McGrail et al. (2015). This threshold was chosen with a policy assumption that tries to minimize the distance that patients travel as a way to reduce contagion risks for others. In the second part of the analysis, we calculated accessibility considering as a threshold the maximum distance to the nearest hospital in each city. This threshold can be interpreted as the minimum necessary distance that guarantees that every person can reach at least one hospital, and is normatively in line with the recommendation from the Ministry of Health (Ministério da Saúde, 2020b) that suspected cases should visit their nearest hospital.

Because this treatment of patients with SARS often requires combined availability of ICU beds and mechanical ventilators, we only considered the joint availability of bed/ventilator. Thus, in the case of a hospital with 30 adult ICU beds but only 20 ventilators, we considered only 20 beds (one ventilator per bed). As a rule, however, there are more ventilators than ICU beds. For ICU beds in field hospitals, we assumed there was at least one ventilator available per bed.

A limitation of this method is that it considered the ICU beds and ventilators that are in use, but these might not be the appropriate models for long use as in COVID-19 cases. Another limitation is that we analyzed the attendance capacity of the public healthcare system focusing only on the number of adult ICU beds and ventilators. Other studies should also consider restrictions imposed by the availability of healthcare professionals to staff ICUs when the data is available. Another limitation of the method is that our analyses are restricted to the population and supply of services within cities. This generates two effects. The first is the tendency to underestimate the level of access to services by people who live near the border between two cities, since they could possibly access hospitals in the neighboring city. The second effect is the underestimation of the demand from people from other neighboring cities who can seek admission to hospitals in the 20 cities analyzed. To minimize this second problem, we applied an adjustment factor for hospital admissions by non-residents (Brasil, 2005) according to the size of the population living in each hexagon, as suggested by Paez et al. (2019). A factor of 1.5, for example, would simulate that the size of the demand for admission to each hospital is 50% higher due to patients living in other cities. In a recent study, Servo, Andrade and Amaral (2019) demonstrated that in 2015 on average 30% of the hospitalizations for medium complexity treatments in cities was from patients living in other cities. Based on data from the SUS Hospital Information System for 2019, we calculated the value of the correction factors for each city (Table 1). Note that the correction factor is less than 1 in Guarulhos and São Gonçalo. This means that these cities export more people looking for healthcare in other cities than they receive from other cities.

**Table 1.** Inpatient care for resident and non-resident in the 20 largest Brazilian cities, 2019.

| City | Total residents inpatient care (A) | Total of inpatient care (B) * | Adjustment factor B / A |
| --- | --- | --- | --- |
| Rio de Janeiro | 226,630 | 283,946 | 1.3 |
| São Paulo | 608,351 | 694,008 | 1.1 |
| Belo Horizonte | 154,761 | 272,452 | 1.8 |
| Curitiba | 126,034 | 166,789 | 1.3 |
| Brasília | 181,564 | 229,614 | 1.3 |
| Fortaleza | 142,076 | 214,470 | 1.5 |
| São Gonçalo | 38,692 | 27,362 | 0.7 |
| Porto Alegre | 104,203 | 183,193 | 1.8 |
| Duque de Caxias | 42,280 | 44,204 | 1.0 |
| Goiânia | 77,235 | 151,762 | 2.0 |
| Campinas | 56,122 | 79,007 | 1.4 |
| Guarulhos | 64,252 | 55,369 | 0.9 |
| Salvador | 154,404 | 237,961 | 1.5 |
| Recife | 107,144 | 298,498 | 2.8 |
| Campo Grande | 52,322 | 68,952 | 1.3 |
| Maceió | 48,439 | 88,008 | 1.8 |
| Belém | 68,667 | 106,579 | 1.6 |
| Manaus | 113,926 | 124,372 | 1.1 |
| São Luís | 60,137 | 105,002 | 1.7 |
| Natal | 45,359 | 93,285 | 2.1 |

Source: Authors own elaboration.
Obs.
- Hospitalizations by place of residence (A), and hospitalizations by hospital location (B). - \* Accounts for hospitalizations of patients who live inside the municipality and patients from outer areas.

## 5. Results

### 5.1 Access to health services

In the 20 largest Brazilian cities nearly 228 thousand low-income people older than 50 years live more than 30 minutes by walking from a health unit that provides triage and referral services to patients with suspected infection (Table 2). The primary health care units and emergency care units, in particular, play a fundamental role as entry points to the system, to avoid hazardous agglomeration of patients with suspected coronavirus infection, so the spatial capillarity and longitudinality of this service are very important.

**Table 2.** Low-income population above 50 years old with access to healthcare in Brazil’s 20 largest cities, 2020.

| City | Total population | Vulnerable population** | (A)<br>Vulnerable Pop.<br>with poor access to<br>basic health<br>services | (B)<br>Vulnerable Pop.<br>with poor access to<br>ICU<br>hospitalization | (B) /<br>Vulnerable<br>Pop.<br>(%) |
| --- | --- | --- | --- | --- | --- |
| Rio de Janeiro | 6592.2 | 692.5 | 51.9 | 384.3 | 55.5 |
| São Paulo | 12142.6 | 1053.6 | 33.2 | 263.1 | 25 |
| Brasília | 3052.5 | 180.3 | 21.1 | 121 | 67.1 |
| Curitiba | 1927 | 172.9 | 5.1 | 116.4 | 67.3 |
| Belo Horizonte | 2469.9 | 244 | 7.2 | 92.3 | 37.8 |
| Fortaleza | 2651.8 | 193.5 | 6.5 | 77.7 | 40.2 |
| São Gonçalo | 1075.4 | 112.9 | 8.8 | 72.6 | 64.3 |
| Duque de Caxias | 905.1 | 81.3 | 13.5 | 67 | 82.4 |
| Porto Alegre | 1480.5 | 159.6 | 8.6 | 60.3 | 37.8 |
| Goiânia | 1509.4 | 118.3 | 11.4 | 59.4 | 50.2 |
| Campinas | 1208.9 | 115.1 | 12.2 | 58.1 | 50.5 |
| Guarulhos | 1389.9 | 98.7 | 4.3 | 48 | 48.6 |
| Recife | 1607 | 147.1 | 0.6 | 42.9 | 29.2 |
| Campo Grande | 895.6 | 69.7 | 5 | 42.9 | 61.5 |
| Maceió | 1042 | 74.2 | 8.4 | 38.2 | 51.5 |
| Salvador | 2831.6 | 217.4 | 7.8 | 35.3 | 16.2 |
| Belém | 1360.1 | 97.2 | 8.7 | 32.9 | 33.8 |
| Manaus | 2216.1 | 111.6 | 2.3 | 24.8 | 22.2 |
| São Luís | 1080.4 | 69.3 | 10.2 | 18.6 | 26.8 |
| Natal | 867.9 | 63 | 1.6 | 10.2 | 16.2 |
| Total | 48305.9 | 4072.2 | 228.4 | 1666 | 40.9 |

**Source:** authors own elaboration using data from Pereira et al. (2019), healthcare facilities data from CNES on February 2020, and population projections for 2020 from Freire et al (2020).
Obs.
- Population in thousands. - \*\* Population above 50 years old and in the bottom half of the income distribution. - (A) Low-income people above 50 years old who cannot access a healthcare facility in less than 30 minutes walking. - (B) Low-income people above 50 years old who live more than 15 Km away from the nearest hospital with an ICU bed and mechanical ventilator.

The highest proportion of people in this situation were found in the cities of Rio de Janeiro, São Paulo, Brasília, Duque de Caxias, Campinas and Goiânia. These six cities concentrate more than 60% of the vulnerable population who live more than 30 minutes on foot from a health service point. However, analysis of the proportion in each city reveals that Duque de Caxias, São Luís, Brasília, Maceió and Campinas stand out with more than 10% of their vulnerable population living more than 30 minutes from an establishment qualified for this triage.

Furthermore, Table 2 also shows there are some 1.6 million vulnerable people who live farther than 5 Km from a health unit able to admit patients in serious condition due to COVID-19. This total represents 41% of the vulnerable population in the 20 cities. These numbers vary widely across cities due to the different patterns of urban occupation and the spatial distribution of healthcare facilities in each area. Cities like Rio de Janeiro, São Paulo, Brasília and Curitiba stand out for having more than 100 thousand inhabitants in potentially vulnerable conditions and with poor access to hospitals with ICU beds and mechanical ventilators. It is also noteworthy that half of the 20 cities have more than 50% of their vulnerable population living farther than 5 Km from inpatient facilities.

As important as estimating how many vulnerable people have poor access to healthcare is mapping where this population lives. Figures 1 to 4 present for four selected cities the size and living place of the low-income population older than 50 years who (A) cannot access any primary healthcare establishment in less than 30 minutes by walking; and (B) live more than 5 Km from the nearest hospital with at least one ICU bed and one ventilator. For the sake of brevity, we only present the maps for Sao Paulo, Rio de Janeiro, Fortaleza and Manaus, the 4 cities most affected by COVID-19 in Brazil. As of the end of June, these four cities alone concentrated 18% of COVID-19 confirmed cases and 32% of deaths in the country. These maps show that vulnerable populations with poor access to health services are mostly located in urban peripheries, indicating those areas which would be good candidates for local policy interventions, such as setting up field hospitals or engaging pre-hospital mobile units (such as the Urgent Mobile Response Service - SAMU) or community health agents.

**Figure 1.**
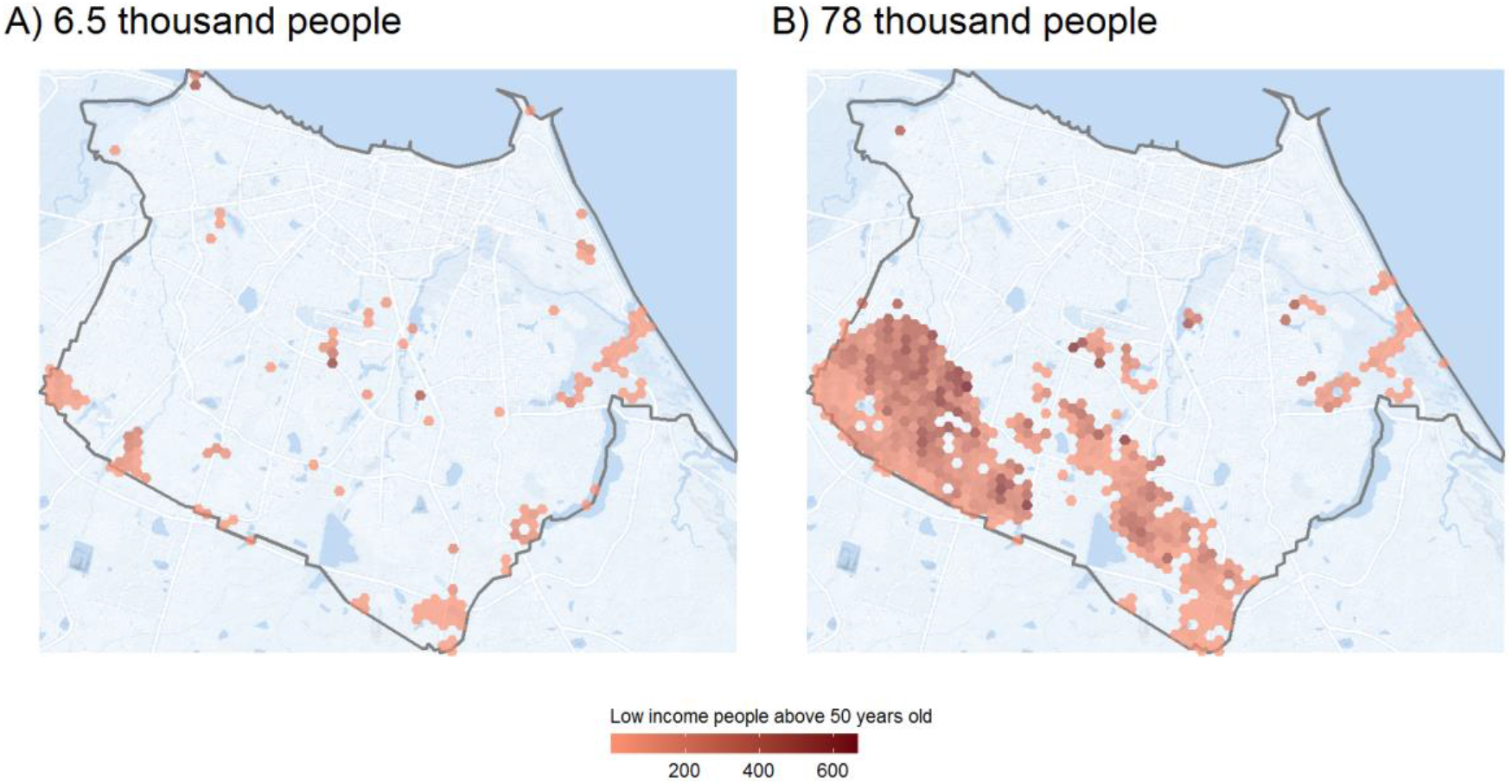
Access to COVID-19 healthcare. Fortaleza, 2020. (A) Vulnerable population who cannot access a basic healthcare facility in less than 30 minutes walking. (B) Vulnerable population who lives farther than 5 Km to the nearest hospital with ICU bed and mechanical ventilator.

**Figure 2.**
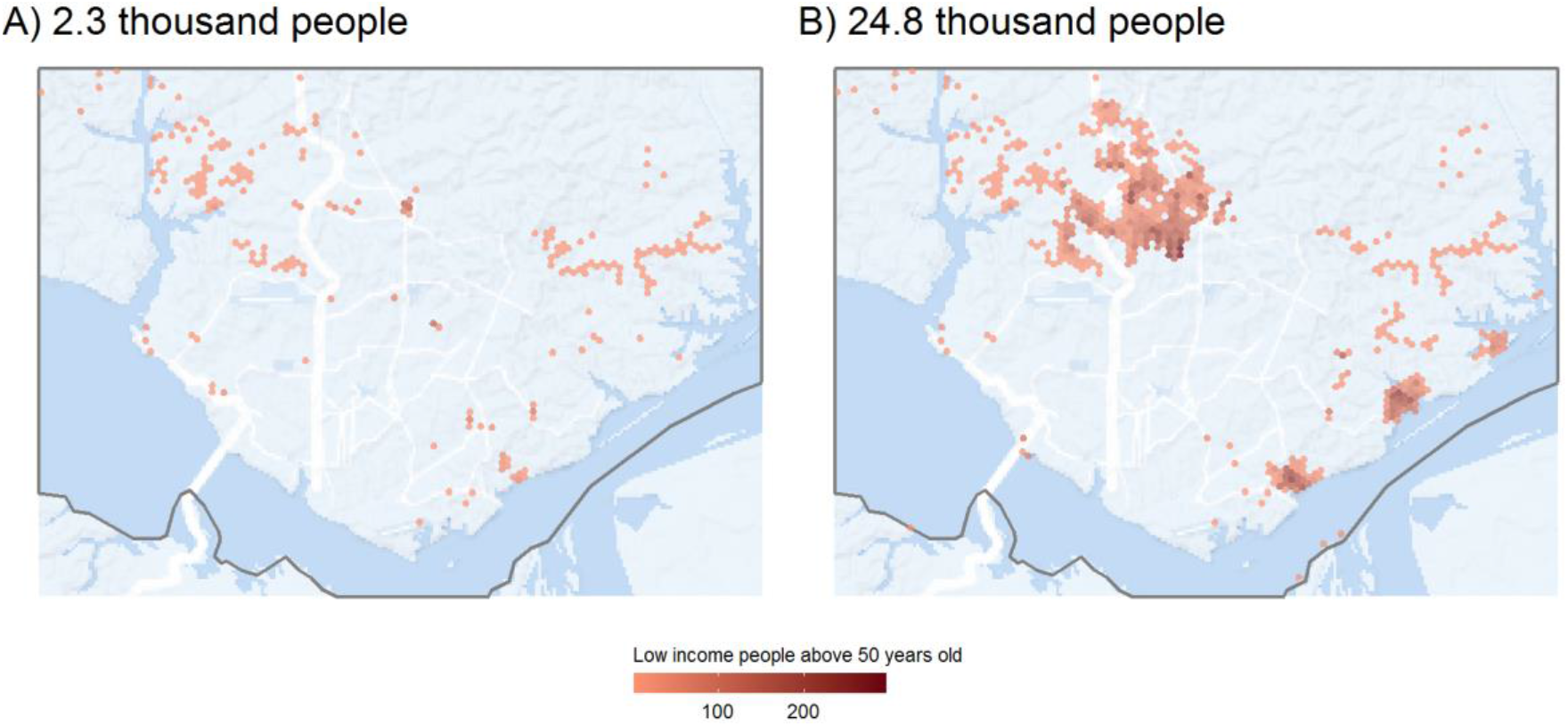
Access to COVID-19 healthcare. Manaus, 2020. (A) Vulnerable population who cannot access a basic healthcare facility in less than 30 minutes walking. (B) Vulnerable population who lives farther than 5 Km to the nearest hospital with ICU bed and mechanical ventilator.

**Figure 3.**
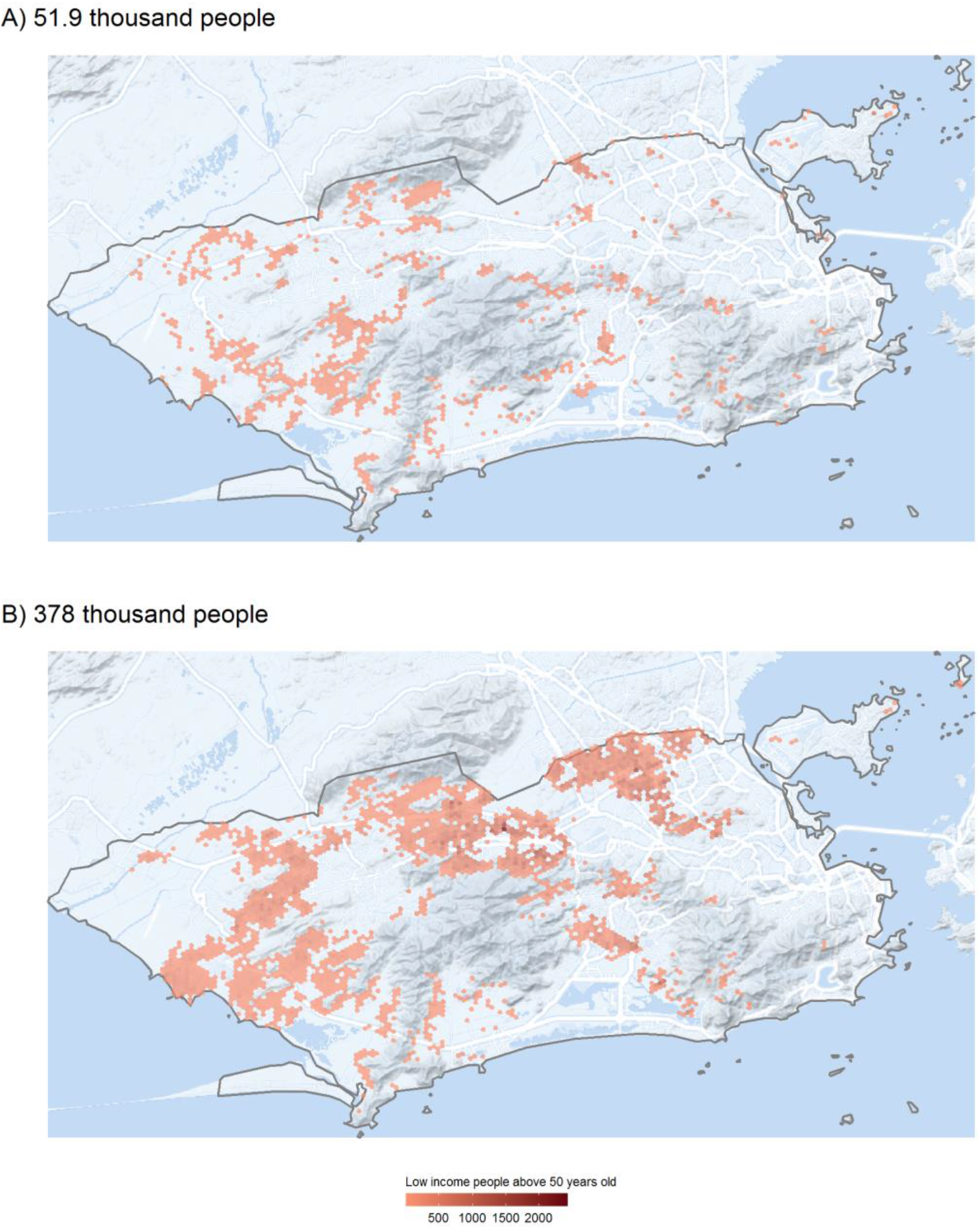
Access to COVID-19 healthcare. Rio de Janeiro, 2020. (A) Vulnerable population who cannot access a basic healthcare facility in less than 30 minutes walking. (B) Vulnerable population who lives farther than 5 Km to the nearest hospital with ICU bed and mechanical ventilator.

**Figure 4.**
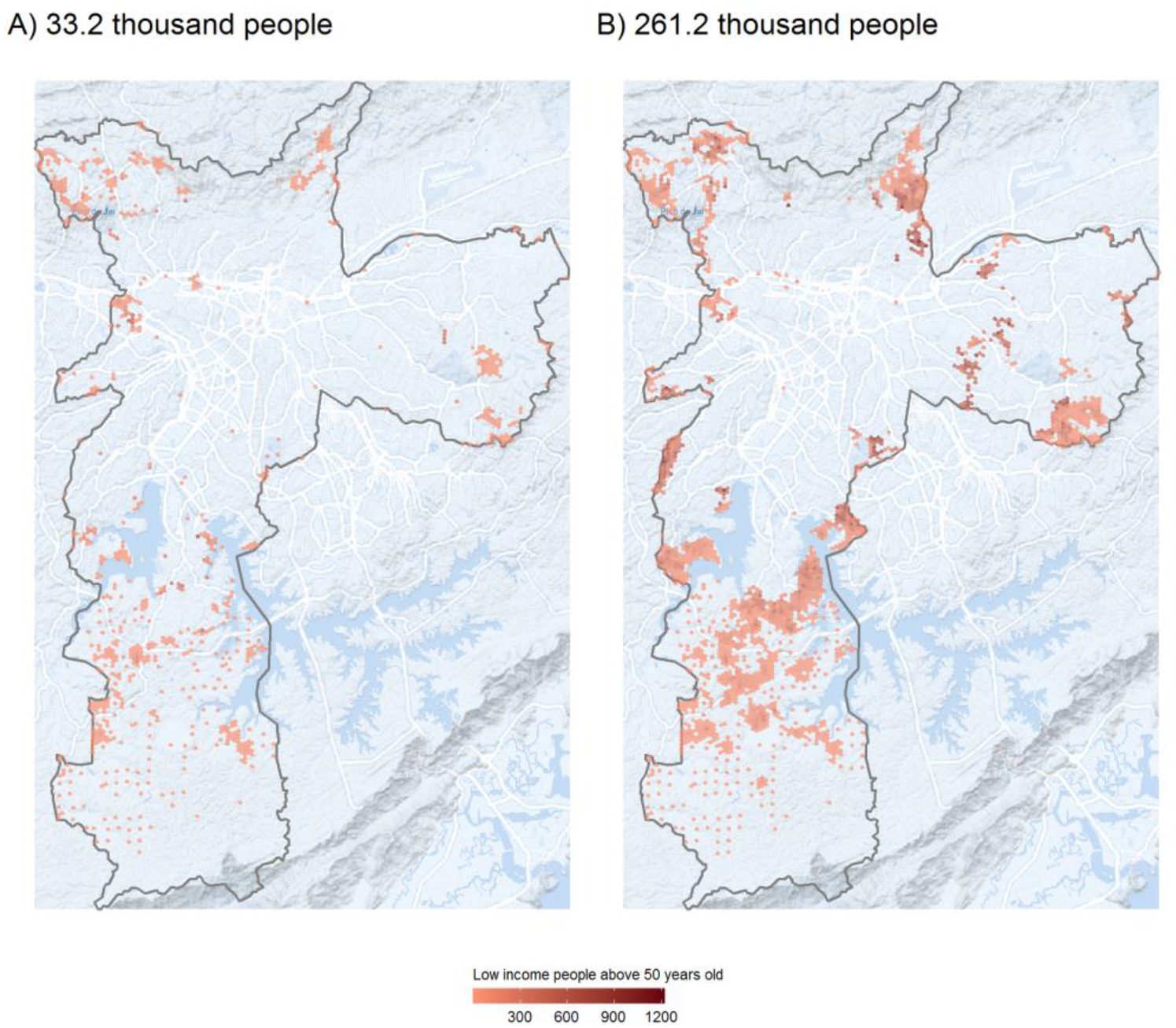
Access to COVID-19 healthcare. São Paulo, 2020. (A) Vulnerable population who cannot access a basic healthcare facility in less than 30 minutes walking. (B) Vulnerable population who lives farther than 5 Km to the nearest hospital with ICU bed and mechanical ventilator.

### 5.2. Health system capacity

A basic indicator to measure the capacity of a health system is the number of hospital beds per inhabitant in a given area. According to the parameters defined by the Brazilian Ministry of Health (Brasil, 2015) the minimum standard provision should be 1 adult ICU bed for each 10 thousand people^4^. The average number of adult ICU beds with ventilators in hospitals in the public health system in the 20 largest Brazilian cities is 1.06 per 10 thousand people (Table 3). This value is only slightly higher than the minimum recommended by the Ministry of Health under normal circumstances. The value of 1.06 can be considered insufficient in an epidemic situation, posing a risk of overload in scenarios for COVID-19 contagion indicated in previous studies (Castro et al., 2020; Noronha et al., 2020). The ratio of beds per population also varies greatly across cities (Table 3). Thirteen out of the 20 cities analyzed are below the recommended level of service and by July 1st most of these cities had bed ICU bed occupancy rates above 80% (Folha de S.Paulo, 2020).

**Table 3.** Number of adult UCI beds and mechanical ventilators in SUS per 10 thousand people in Brazil’s 20 greatest municipalities, 2020.

| **Municipality** | **ICU beds\*** | **Population (in thousands)\*\*** | **ICU beds per 10 thousand people** |
| --- | --- | --- | --- |
| São Gonçalo | 149 | 760.5 | 2 |
| Goiânia | 596 | 2965.9 | 2 |
| Belo Horizonte | 792 | 4348.2 | 1.8 |
| Rio de Janeiro | 1419 | 8259.4 | 1.7 |
| Porto Alegre | 327 | 2602.8 | 1.3 |
| Salvador | 565 | 4363.9 | 1.3 |
| São Paulo | 1211 | 13852.3 | 0.9 |
| Campo Grande | 102 | 1180.3 | 0.9 |
| Curitiba | 211 | 2550.1 | 0.8 |
| Campinas | 141 | 1701.9 | 0.8 |
| Guarulhos | 93 | 1197.7 | 0.8 |
| Recife | 373 | 4477 | 0.8 |
| São Luís | 159 | 1886.4 | 0.8 |
| Belém | 139 | 2111 | 0.7 |
| Manaus | 170 | 2419.3 | 0.7 |
| Natal | 123 | 1784.9 | 0.7 |
| Fortaleza | 241 | 4003 | 0.6 |
| Brasília | 181 | 3860.3 | 0.5 |
| Maceió | 82 | 1893.2 | 0.4 |
| Duque de Caxias | 30 | 946.3 | 0.3 |
| Total | 7,104 | 67,164.4 | 1.06 |

Source: authors own elaboration using data from Pereira et al. (2019), healthcare facilities data from CNES on February 2020, and population projections for 2020 from Freire et al (2020). obs.: \* Number of ICU beds with mechanical ventilators in the public healthcare system. \*\* City population corrected by the adjustment factor from Table 1.

The greatest variations in the availability of health services, however, occur within cities. In Figures 5 to 8 below we present a set of maps that together give a detailed view of the spatial distribution of hospitals with adult ICU beds and ventilators and the level of access to these services considering competition effects. The maps on **Panel A** show the spatial distribution of hospitals, where each hospital is represented by a circle whose size reflects the ratio between the number of beds/ventilators of that hospital and the population of its catchment area. Although the situations vary across cities, as a rule, downtown areas generally concentrate the greatest number of hospitals, especially the ones with more beds per inhabitant. This is the case for example in Fortaleza, Manaus and Rio de Janeiro. The availability of ICU beds and ventilators to serve patients with severe COVID-19 tends to be considerably lower in the peripheral regions of these cities. In these regions, it is common to observe ratios of ICU beds per 10 thousand inhabitants between 0.5 and1.0. These ratios can be considered critical against a backdrop of an epidemic with growing numbers of patients needing hospitalization for respiratory complications. The example of cities like Rio de Janeiro also illustrate how setting up field hospitals in farther areas can increase the capillarity of health services in epidemic situations like the COVID-19 outbreak.

**Figure 5.**
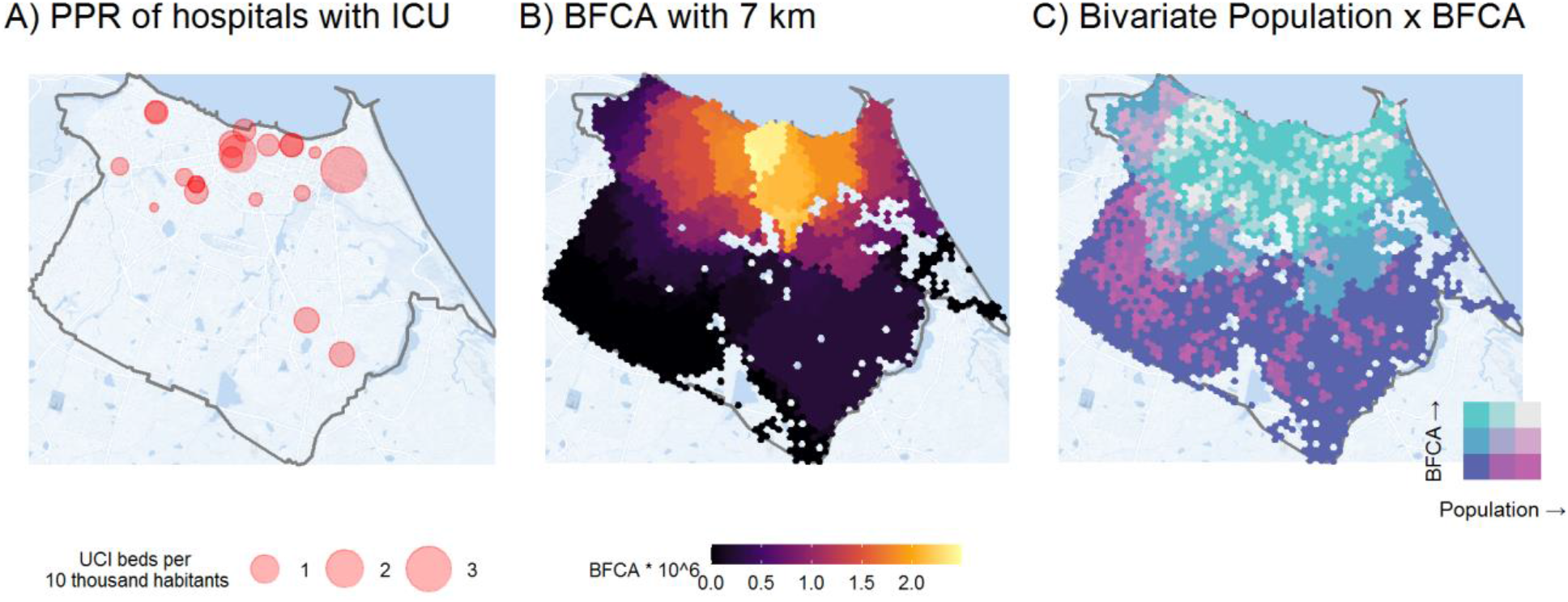
Spatial distribution of hospitals with adult ICU beds and ventilators and the level of access to these services considering competition effects. Fortaleza, 2020.

**Figure 6.**
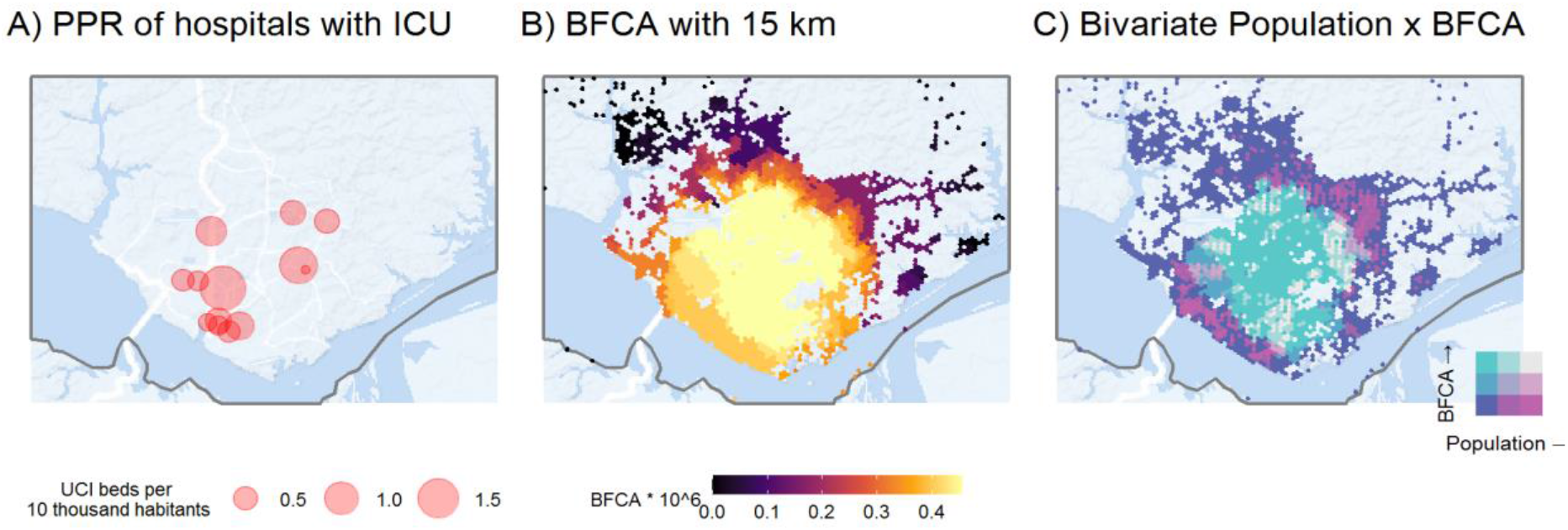
Spatial distribution of hospitals with adult ICU beds and ventilators and the level of access to these services considering competition effects. Manaus, 2020.

**Figure 7.**
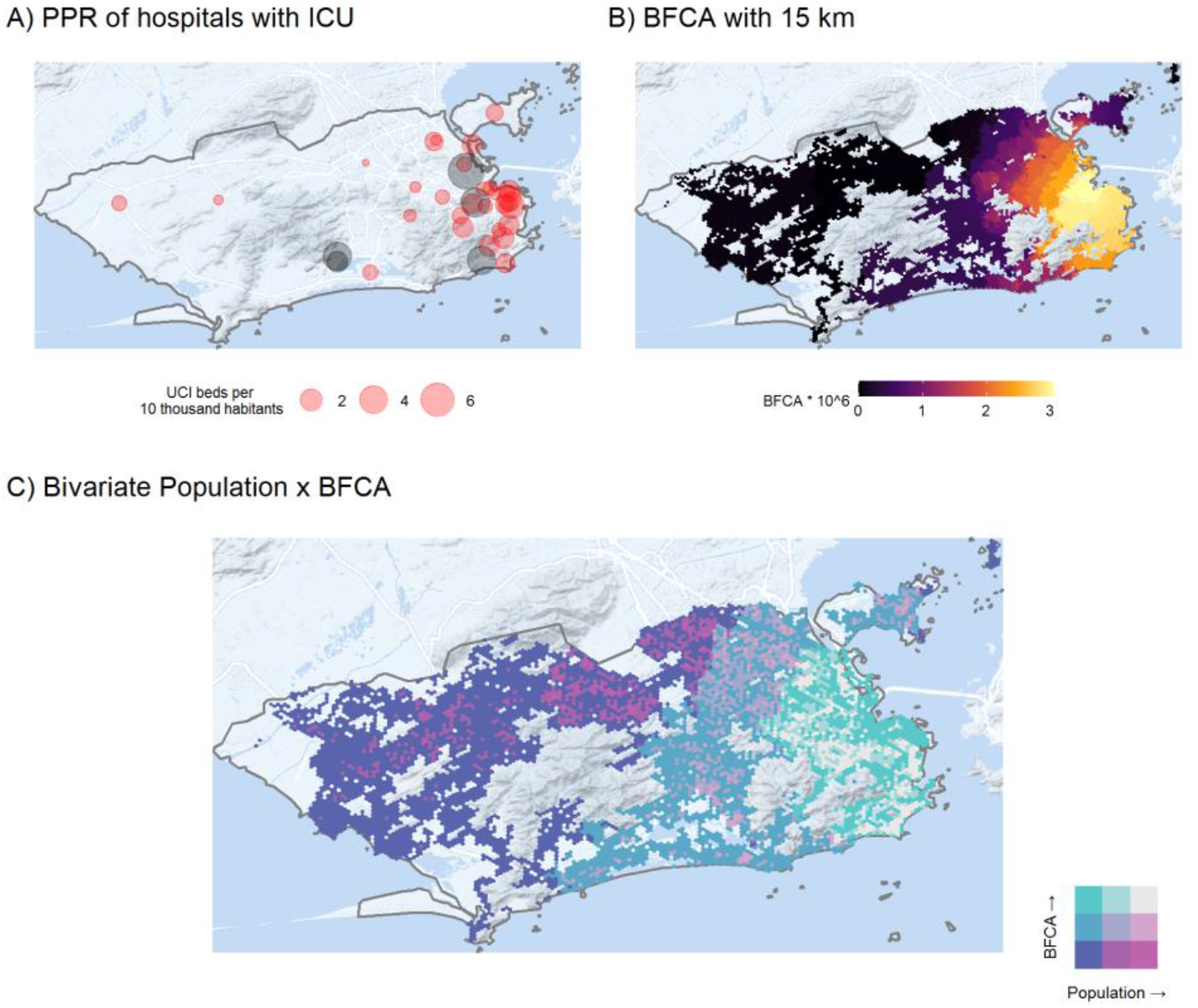
Spatial distribution of hospitals with adult ICU beds and ventilators and the level of access to these services considering competition effects. Rio de Janeiro, 2020. Obs. Gray circles represent field hospitals or temporarily reactivated facilities.

**Figure 8.**
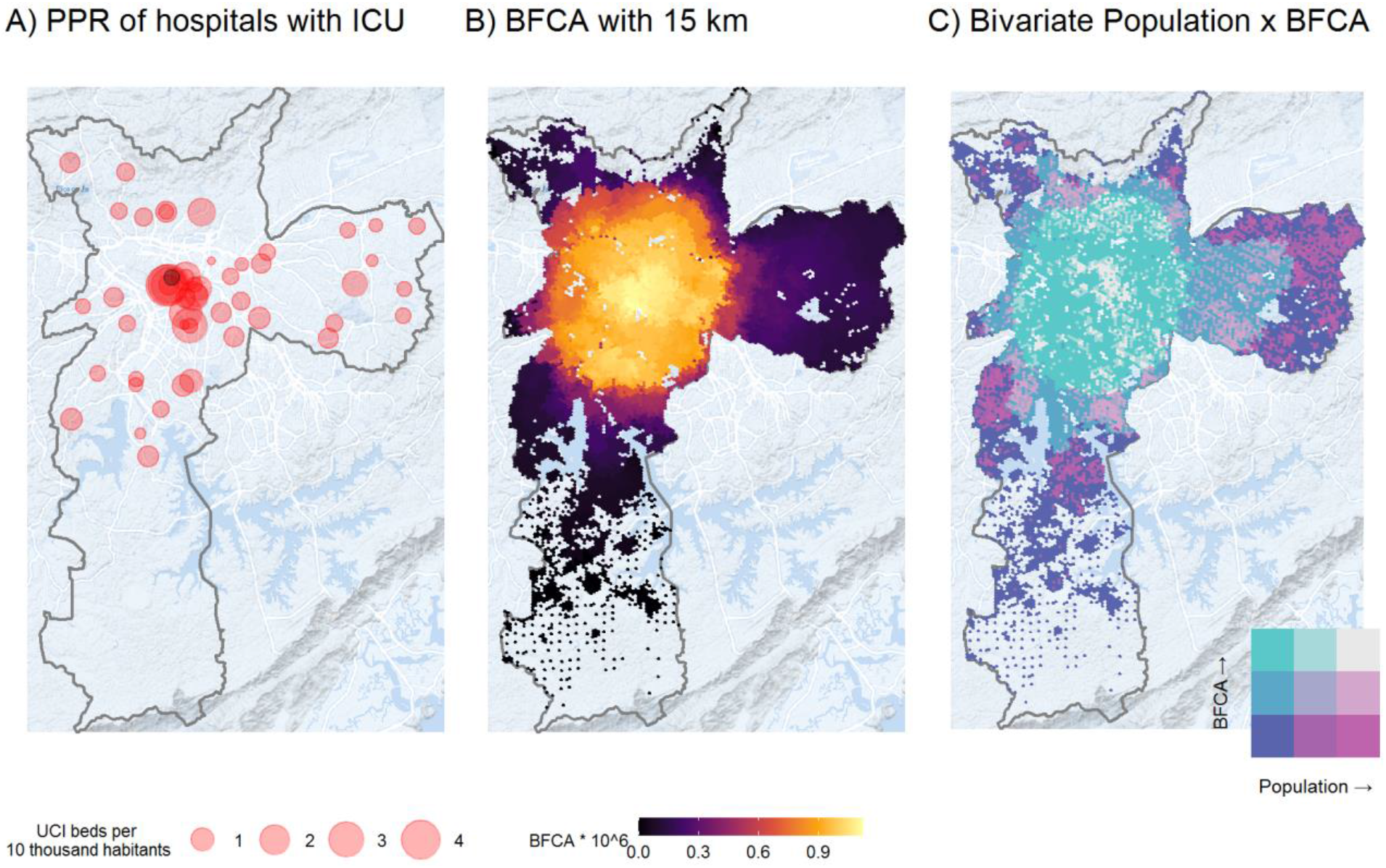
Spatial distribution of hospitals with adult ICU beds and ventilators and the level of access to these services considering competition effects. São Paulo, 2020. Obs. Gray circles represent field hospitals or temporarily reactivated facilities.

Meanwhile, the maps on **Panel B** show the number of ICU beds and ventilators accessible from each location considering at the same time the level of service availability and the potential overlap of demand and supply competition effects estimated with the balanced float catchment area (BFCA). Compared to the results in Panel A, these maps present in more detail how the geographic access to COVID-19 healthcare is particularly higher in central urban areas. This is perhaps more clearly seen in the city of São Paulo, where accessibility to equipped beds steadily declines from the central parts of the city to the periphery. Considering the average population per tile of the hexagonal grid in São Paulo, this indicates that in the regions with the highest accessibility, there are approximately 0.000012 beds per 10,000 people serving on average 1,240 persons. This translates into 9.76e-6 beds per person, compared to 8.74e-5 beds per person average for São Paulo. This shows how the regional PPR can be misleading, by assuming that every person in the region has equal access to medical facilities. Analysis using the BFCA indicator also illustrates how living close to a hospital does not necessarily translate into high levels of accessibility, once congestion effects are taken into account. This is the case in cities like Fortaleza, Rio de Janeiro, and São Paulo, where some peripheral neighborhoods, despite being close to a hospital, face poor access to health services due to the limited capacity of the healthcare system to support the expected demand of much larger areas.

Finally, the maps on **Panel C** present bivariate choropleth maps with the combined spatial distribution of population densities and accessibility levels by automobile using city-specific thresholds so that every person could reach at least one hospital. This panel complements previous figures by highlighting those areas with large populations underserved by healthcare services (bright pink = larger population and lower accessibility) and those areas which face higher service levels for a comparable lower demand (bright green = smaller population but high accessibility). It is possible to see that even in those places with low accessibility, there are pockets where the situation is made worse by afflicting larger populations. The figures in Panel C are useful to differentiate low-accessibility areas with large and small population numbers, which can provide actionable information for policy makers to choose which low-accessibility areas should be prioritized in emergency situations.

The geographic access to COVID-19 healthcare in Brazil presents not only spatial but also marked social inequalities. Figure 9 shows the magnitude of the racial and income inequalities in access to ICU beds and ventilators considering competition effects. One of the most extreme cases is the capital of the country Brasília, where the number of ICU beds with ventilators accessible by the wealthiest population is more than 6 times larger than for the poor. While racial inequalities are relatively lower compared to inequalities by income, they are still present in most cities. This is particularly true in Brasília, São Paulo and Belo Horizonte, where black communities can only access half as many health resources as the white population.

**Figure 9.**
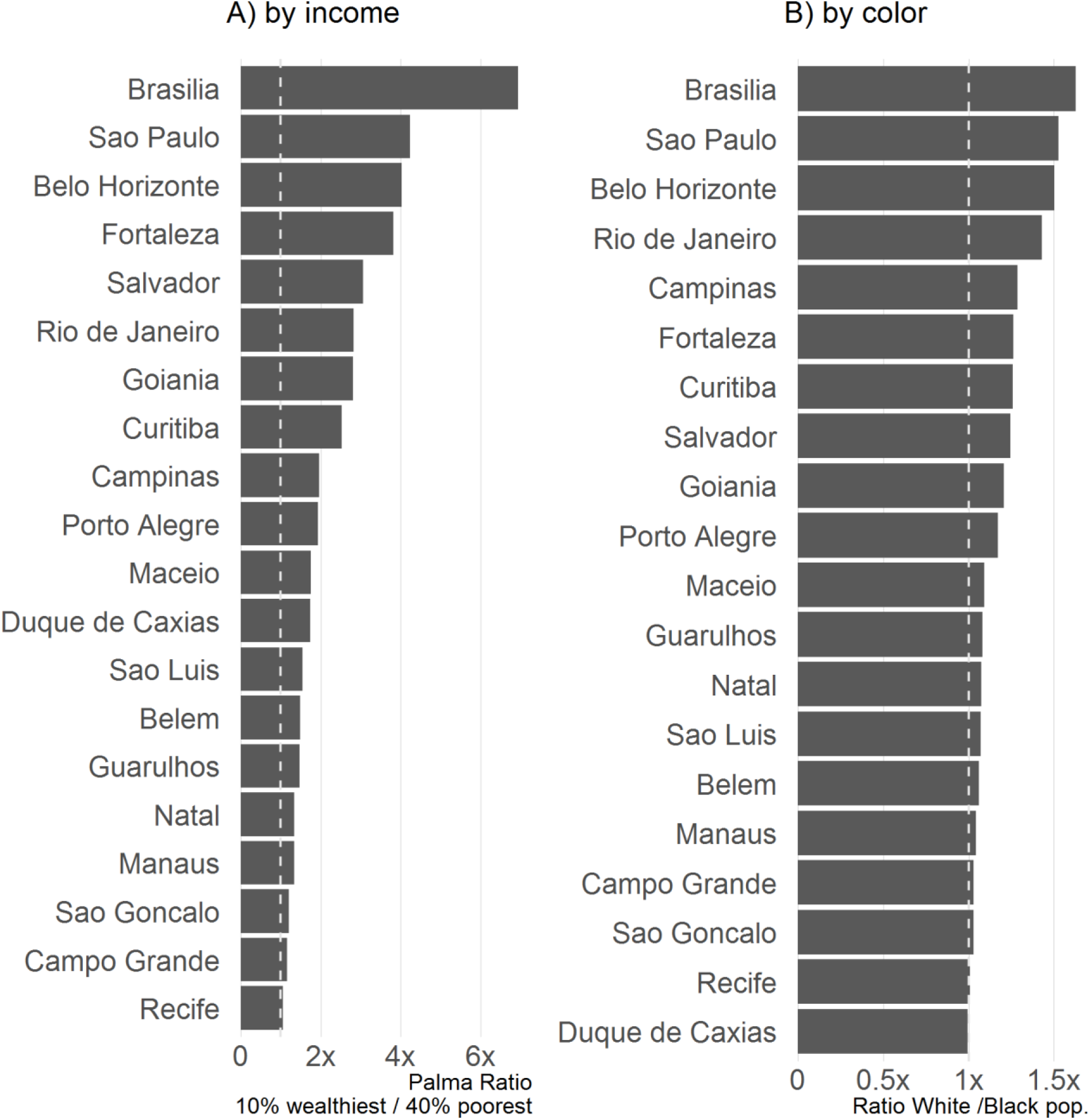
Income and racial inequalities in access to ICU beds and ventilators considering competition effects. Brazil’s 20 largest cities, 2020.

In summary, our findings point to a worrying pattern. Across Brazil’s 20 largest cities, we find substantially lower healthcare system capacity in urban peripheral areas and among low-income and black communities. In particular, urban peripheries with high population density coupled with low incomes and poor sanitation services create a worrying scenario with strong potential for propagation of the COVID-19 precisely among communities that are most vulnerable to the disease and with lowest access to healthcare. Combined, the analyses presented provide valuable information that can help local authorities map the areas which should receive more immediate response from local healthcare community agents and perhaps the construction of new field hospitals to address short term needs induced by the COVID-19 pandemic.

## 6. Final remarks

This paper examined the geographical access to COVID-19 healthcare in the 20 largest Brazilian cities, looking at both healthcare facilities with capacity for triage and referral of patients with suspected COVID-19 infections to hospitals, as well as those able to treat patients in serious condition through the support of ICU beds and mechanical ventilators. We mapped approximately 230 thousand vulnerable people (low-income above 50 years old) living more than 30 minutes walking from a public healthcare facility able to test for COVID-19 or refer suspected cases to hospitals. Our results also indicate that some 1.6 million vulnerable people live farther than 5 Km from a hospital with capacity for ICU admission. Because patients suspected with COVID-19 might face mobility constraints due to grave conditions, it becomes crucial to develop strategies for provision of transport and health services to these people. This is particularly true of low-income people living on the outskirts of cities, where there are fewer mobility options and where health services are scarcer.

The study also analyzed the support capacity of the public health systems in the largest Brazilian cities, looking at the number of ICU beds/ventilators per person in the catchment area of each hospital and accessible by the population. We find that thirteen out of the 20 cities analyzed have fewer ICU beds with ventilators than the minimum level recommended by the Health Ministry, which could be insufficient to deal with large growth of demand for hospital admissions in the most optimistic scenarios of COVID-19 propagation in Brazil. Accessibility analysis using the new balanced float catchment area (BFCA) shows this scenario is particularly worrisome when we account for competition effects on both supply and demand for health services. The BFCA estimates show substantially lower access to health services in low-income and black communities in urban peripheries, which generally face scarcer supply of health equipment and could more easily be overwhelmed by the near-future hospitalization demands.

As a whole, the study illustrates how transport accessibility analyses can provide actionable information to help local governments improve access to healthcare services during pandemic outbreaks. Combined, the analyses in the paper put disadvantaged communities with poor access to health services on the map, indicating in which neighborhoods local authorities could prioritize to build makeshift hospitals or engage mobile units or health community agents. These analyses also help local authorities identify which hospitals that might face the greatest admission overload due to competition effects, and hence would need supplementary funding to expand capacity. The application of the novel BFCA in this paper illustrates how considering competition effects in access to healthcare can have important but often overlooked implications for policy planning.

Future research could potentially combine the results of this paper with scenarios for COVID-19 infection and hospitalization rates to generate estimates of demand for beds and ventilators on an intra-urban scale. Further analysis could also indicate the areas where the construction of new makeshift hospitals would be more effective to improve healthcare accessibility at the city level and for vulnerable groups in particular. More studies however are still needed to investigate how the availability of healthcare professionals could hinder the population access to healthcare services and about the potential role of community health agents in serving people in more remote areas.

Finally, although the reduction of public transportation services can potentially reduce urban mobility levels and limit the dissemination of the virus, it can also restrict access to healthcare facilities by healthcare professionals along with low-income patients and their families, who have few mobility options. The reduction of transportation services without a coordinated restriction of other activities causes longer waiting periods, and can consequently increase the agglomeration of people at transport stations and crowding aboard vehicles, aggravating dissemination of the virus. In this respect, it is important for local managers to reorganize the public transit service to assure better access to healthcare facilities. This could include provision of exclusive services to healthcare professionals and providers of other essential services, without impairing the supply of services of regular lines.

## Data Availability

Data is available upon request

https://www.ipea.gov.br/acessooportunidades/en/

## Footnotes

1 More information about the Access to Opportunities Project and its databases are available at: https://www.ipea.gov.br/acessooportunidades/en/.

2 Ideally, it would be important also to consider the population with comorbidities, such as hypertension, diabetes and cardiovascular or respiratory diseases, because people with these profiles are at greater risk of COVID-19 infection, with greater severity and lethality (Guan et al., 2020; Yang et al., 2020). However, data with this level of detail are not yet available.

3 The following types of healthcare units for first-response services and triage were considered: community health posts, basic health centers/units, polyclinics, general hospitals, specialized hospitals, mixed care units, general first aid posts, specialized first aid posts, and units for indigenous health response. As established by the Management Protocol for the New Coronavirus (*Protocolo de Manejo para o Novo Coronavirus*), “All patients who seek health services (Primary Health Response Units, Emergency Care Units, First Aid Posts, Mobile Pre-Hospital Service Units and Hospitals), must be submitted to clinical triage that includes early recognition of suspected cases, and if necessary, immediate referral of the patient to an area separated from those that contain respiratory and hand hygiene supplies” (free translation from Ministério da Saúde, 2020b).

4 Edict 1,101 of June 12, 2002, and Resolution 7 of February 24, 2010.

## Notes

### Competing Interest Statement

The authors have declared no competing interest.

### Funding Statement

No external funding was received

### Author Declarations

The study is based on computational models and did not involve research on human subjects

